# Longitudinal associations between mild behavioral impairment, sleep disturbance, and progression to dementia

**DOI:** 10.1101/2023.09.24.23296033

**Authors:** Dinithi Mudalige, Dylan X. Guan, Maryam Ghahremani, Zahinoor Ismail

**Affiliations:** University of Calgary, Calgary, AB, Canada; Hotchkiss Brain Institute, University of Calgary, Calgary, AB, Canada; Department of Psychiatry, Department of Community Health Sciences, Department of Clinical Neurosciences, Hotchkiss Brain Institute, University of Calgary, Calgary, AB, Canada; Department of Clinical and Biomedical Sciences, Faculty of Health and Life Sciences, University of Exeter, Exeter, UK

**Author notes:** Corresponding Author: Zahinoor Ismail 3280 Hospital Dr. NW Calgary AB Canada T2N 4Z6.

**Keywords:** Alzheimer’s disease, sleep disorders, neurobehavioral manifestations, dementia, longitudinal studies

## Abstract

**Background:** Clinical guidelines recommend incorporating non-cognitive markers like mild behavioral impairment (MBI) and sleep disturbance (SD) into dementia screening to improve detection.

**Objective:** We investigated the longitudinal associations between MBI, SD, and incident dementia.

**Methods:** Participant data were from the National Alzheimer’s Coordinating Center in the United States. MBI was derived from the Neuropsychiatric Inventory Questionnaire (NPI-Q) using a published algorithm. SD was determined using the NPI-Q nighttime behaviors item. Cox proportional hazard regressions with time-dependant variables for MBI, SD, and cognitive diagnosis were used to model associations between baseline 1) MBI and incident SD (n=11277); 2) SD and incident MBI (n=10535); 3) MBI with concurrent SD and incident dementia (n=13544); and 4) MBI without concurrent SD and incident dementia (n=11921). Models were adjusted for first-visit age, sex, education, cognitive diagnosis, race, and for multiple comparisons using the Benjamini-Hochberg method.

**Results:** The rate of developing SD was 3.1-fold higher in older adults with MBI at baseline compared to those without MBI (95%CI: 2.8-3.3). The rate of developing MBI was 1.5-fold higher in older adults with baseline SD than those without SD (95%CI: 1.3-1.8). The rate of developing dementia was 2.2-fold greater in older adults with both MBI and SD, as opposed to SD alone (95%CI:1.9-2.6).

**Conclusions:** There is a bidirectional relationship between MBI and SD. Older adults with SD develop dementia at higher rates when co-occurring with MBI. Future studies should explore the mechanisms underlying these relationships, and dementia screening may be improved by assessing for both MBI and SD.

## INTRODUCTION

Dementia is currently the seventh leading cause of death worldwide, and 152 million people are expected to develop dementia by 2050 [1, 2]. Alzheimer’s disease (AD) is the most common cause of dementia [3]. With some recent exceptions, clinical trials for AD disease-modifying therapies have largely been unsuccessful at meeting primary endpoints, partly because of administration too late in the disease course [4]. Therefore, there is a crucial need for improved detection of older adults at risk of dementia to facilitate the development of therapeutic and preventative interventions and to identify those with early-stage disease potentially suitable to treat with disease-modifying therapies.

Clinical guidelines have recommended the use of non-cognitive markers of dementia for improved dementia detection [5, 6]. Two of these markers are mild behavioral impairment (MBI) and sleep disturbance (SD). MBI is a neurobehavioral syndrome that captures later-life emergent and persistent neuropsychiatric symptoms (NPS), representing a change from longstanding behavior or personality, as a high-risk state for incident dementia [7]. MBI symptoms are categorized into five domains: decreased drive and motivation (apathy), affective dysregulation (mood and anxiety symptoms), impulse dyscontrol (agitation, impulsivity, abnormal reward salience), social inappropriateness (impaired social cognition), and abnormal perception or thought content (hallucinations and delusions, i.e., psychotic symptoms). SD can include a variety of disturbances, including insomnia, REM sleep disorder, sleep apnea, and restless leg syndrome, the most common of which is insomnia (i.e., difficulties falling and staying asleep), resulting in excessive daytime sedation [8].

Cross-sectional studies have indicated that MBI and SD may be associated with each other. For instance, behavioral symptoms may contribute to insomnia [9]. Conversely, SD may increase susceptibility to behavioral symptoms [10, 11].

Both MBI and SD are associated with greater cognitive decline and dementia. First, studies have indicated that MBI is associated with cognitive impairment cross-sectionally [12, 13] and incident cognitive decline and dementia longitudinally in cognitively normal (CN) older adults [14, 15], mixed CN and mild cognitive impairment (MCI) samples [16–22] and in MCI [23]. Analogous to MBI, several studies have demonstrated associations between SD and cognitive decline leading to dementia [24–26]. This risk extends to the preclinical stage in CN older adults, as both MBI and SD can precede typical symptoms of AD, such as memory loss, and associate with a greater risk of incident dementia [15, 27]. Despite evidence suggesting that both MBI and SD are associated with dementia risk, the relationship between MBI and SD and their combined prognostic utility is poorly understood.

This study investigated bidirectional associations between MBI and SD and determined if the prognostic utility of SD for incident dementia could be improved by further adding MBI to the modeling. We hypothesized that older adults with MBI would develop SD at a faster rate, and vice versa, due to potential positive feedback. Additionally, we hypothesized that sleep-disturbed older adults with MBI would develop dementia at a faster rate than SD older adults without MBI due to the additive effects of MBI and SD.

## METHOD

### Study Design

The National Alzheimer’s Coordinating Center (NACC) Uniform Data Set (UDS) was used, including participant data collected approximately annually from June 2005 to May 2022. The NACC was established by the National Institute on Aging and consisted of data prospectively collected from 45 NIA-funded Alzheimer’s Disease Research Centers (ADRCs) across 26 states since 1999 [28]. Data are primarily collected by trained clinicians and clinical staff from participants and informants. Extensive descriptions of NACC recruitment and data collection procedures have been published elsewhere [28–31]. Participants and informants provided informed consent at the ADRC, and the respective Institutional Review Board approved all studies conducted at ADRCs that provided data to NACC.

### Participants

The overall sample from the NACC-UDS consisted of 45100 participants. For this study to satisfy the MBI criterion of later-life emergence of NPS [7], only participants older than 50 years old (n=1069) with no prior psychiatric or developmental disorders (e.g., post-traumatic stress disorder (PTSD), bipolar disorder, anxiety disorder, schizophrenia, obsessive-compulsive disorder (OCD), Downs syndrome, Huntington’s, or Parkinson’s disease were included (*n* = 14556). Participants missing data for demographic information (n=244) and Neuropsychiatric Inventory Questionnaire (NPI-Q) domains required to derive MBI (*n* = 910) and SD status (n=72) were excluded [Figure 1]. All participants were required to have no diagnosis of dementia at baseline and have at least one follow-up visit. The clinician diagnosis in the NACC-UDS was used to determine cognitive status, including dementia.

**Figure 1.**
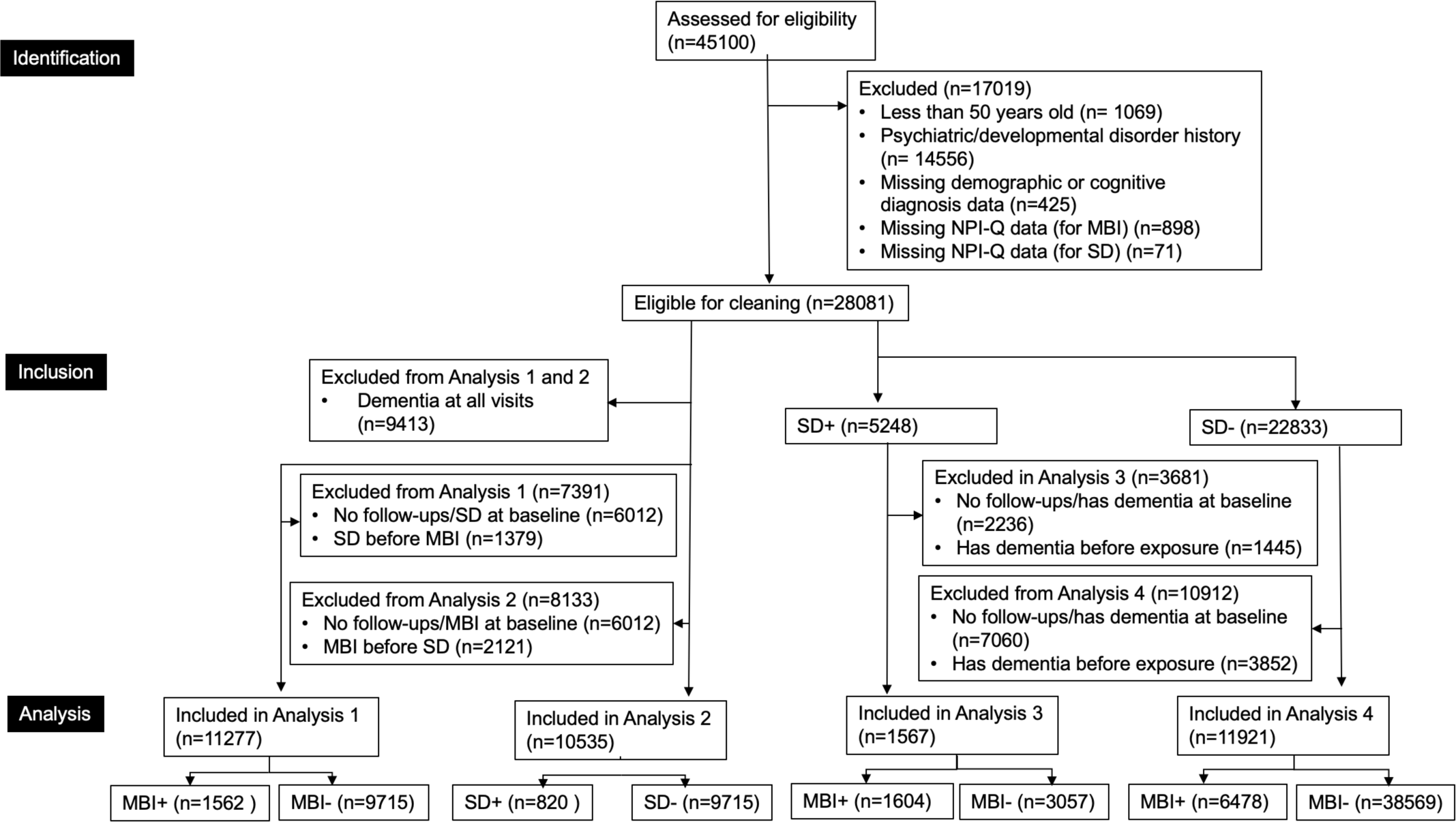
Participant Flow Diagram. Analysis 1 examined whether baseline MBI was associated with incident sleep disturbance. Analysis 2 examined whether baseline sleep disturbance was associated with incident MBI. Analysis 3 explored whether baseline MBI was associated with dementia in those with sleep disturbance. Analysis 4 explored whether baseline MBI was associated with dementia in those without sleep disturbance. Abbreviations: NPI-Q, Neuropsychiatric Inventory Questionnaire; MBI, mild behavioral impairment; SD, sleep disturbance

For the longitudinal analyses examining the association between baseline MBI status and incident SD (and vice versa), additional criteria were applied, as illustrated in Figure 1. The sample sizes for the analyses examining the association between baseline MBI and incident SD and baseline SD and incident MBI were 11277 and 10535, respectively. The sample sizes for the analyses examining the association between baseline MBI within sleep-disturbed older adults and incident dementia and within non-sleep-disturbed older adults and incident dementia were 13544 and 11921, respectively.

### Measures

#### Sleep disturbance

SD was measured using the NPI-Q nighttime behaviors item, as described in previous studies [32–35]. The nighttime behaviors item asks informants: “Does the patient awaken you during the night, rise too early in the morning, or take excessive naps during the day?” to which informants respond “Yes” or “No” and rate the severity of the SD on a scale of 1-3 if present. For the present study, only the reported presence or absence of SD was utilized.

#### Mild Behavioral Impairment

MBI was derived from the NPI-Q [36] using a published algorithm [37, 38]. The MBI decreased motivation score, with a range of 0-3, was directly derived from the NPI-Q apathy score. The MBI emotional dysregulation score, with a range of 0-9, was derived by summing the NPI-Q depression, anxiety, and elation scores. The MBI impulse dyscontrol score, with a range of 0-9, was derived by summing the NPI-Q agitation, irritability, and motor behavior scores. The MBI social inappropriateness score, with a range of 0-3, was derived directly from the NPI-Q disinhibition score. Lastly, the MBI abnormal perception/thought content score, ranging from 0-6, was derived by summing the NPI-Q delusions and hallucination scores. The global MBI score was then derived by summing up the five domain scores for each visit, with a range of 0-30, with higher scores indicating greater MBI symptom severity. However, for this study, MBI was dichotomized, where a global MBI score >0 indicated presence. To meet the MBI symptom persistence criterion, symptoms had to be present (global MBI score>0) at two consecutive NACC visits prior to dementia onset [7].

### Statistical Analysis

Participant demographics were summarized using descriptive statistics (means, standard deviations, medians, interquartile ranges, ranges, counts, and percentages). To compare between groups, Wilcoxon signed-rank tests, and chi-squared tests were used.

To determine baselines for survival models, a participant-specific baseline was generated based on the first instance of symptom positivity for both MBI and SD. For each participant, the baseline was set to the visit at which symptoms of either MBI or SD were endorsed to correspond to the onset of behavioral symptoms or sleep disturbance. For participants who did not develop MBI or SD before dementia incidence, the first study visit was considered baseline. MBI positivity required two consecutive NPS+ visits consistent with the MBI symptom persistence criterion [7]. The presence of SD, however, required only a single-visit endorsement of symptoms.

Survival analyses were completed utilizing Kaplan-Meier (KM) survival curves and Cox proportional hazards regressions with time-dependent variables for MBI, SD, and cognitive diagnosis. Four models were built for 1) the association between baseline MBI status and incident SD; 2) the association between baseline SD and incident MBI; 3) the association between baseline MBI with concurrent SD and incident dementia; 4) the association between baseline MBI without concurrent SD and incident dementia. Participants were followed up for a maximum of 16.3 years for analyses 1, 2, and 4 and 15.4 years for the third analysis; however, for ease of visualization, KM curves were truncated to 12 years without omitting any data [39]. Participant age at the first visit, sex, years of education, cognitive diagnosis (CN or MCI), and race were included as covariates. These covariates were chosen because they are associated with an increased risk for dementia [23, 40, 41]. All p-values were adjusted for multiple comparisons using the Benjamini–Hochberg method to generate q-values. The proportional hazards (PH) assumption of the Cox PH models was confirmed using Schoenfeld residuals. R version 4.0.5 was used to conduct all statistical analyses [42].

## RESULTS

### Participant Characteristics

Baseline participant demographics have been summarized in Table 1. Table 2 include participant characteristics for each model stratified by baseline MBI or SD status. In general, the four analyses had similar sample sizes, with the analysis of baseline MBI status in those without SD and incident dementia having the largest sample of 11921 participants. Across analyses, at baseline, participants were, on average, about 72 years old, had 15 years of education, and were majority female and CN (Table 1).

**Table 1.** Participant characteristics.

| **Table 1. Participant characteristics** |  |  |  |  |
| --- | --- | --- | --- | --- |
|  | **Analysis 1\*** | **Analysis 2\*** | **Analysis 3\*** | **Analysis 4\*** |
| **n** | 11277 | 10535 | 1567 | 11921 |
| **First-Visit Age (Years)** | 72.7 (8.8), 50.0-104.0 | 72.7 (8.8), 50.0-101.0 | 72.2 (8.8), 50.0-101 | 72.9 (8.8), 50.0-104 |
| **Females** | 6658 (59.0) | 6412 (60.9) | 785 (50.1) | 6991 (58.6) |
| **Education (Years)** | 15.7 (3.1), 0-29.0 | 15.7 (3.2), 0-29.0 | 15.4 (3.3), 0-30.0 | 15.7 (3.1), 0-29.0 |
| **Cognitive Status** |  |  |  |  |
| CN | 8579 (76.1) | 8375 (79.5) | 797 (50.9) | 8628 (72.4) |
| MCI | 2698 (23.9) | 2160 (20.5) | 770 (49.1) | 3293 (27.6) |
| **Race: White** | 8721 (77.3) | 8063 (76.5) | 1260 (80.4) | 9269 (77.8) |
| **Predictor Characteristics** |  |  |  |  |
| MBI | 1562 (13.9) | - | 707 (45.1) | 1902 (16.0) |
| No MBI | 9715 (86.1) | - | 860 (54.9) | 10019 (84.0) |
| SD | - | 820 (7.8) | - | - |
| No SD | - | 9715 (92.2) | - | - |

*Note.* \*Analysis 1: Baseline MBI and incident sleep disturbance, Analysis 2: Baseline sleep disturbance and incident MBI, Analysis 3: Baseline MBI and incident dementia within those with sleep disturbance. Analysis 4: baseline MBI and incident dementia in those without sleep disturbance. All values for continuous variables are shown in mean (SD), range. Values were rounded to one decimal place where appropriate. Categorical variables are shown as n (%). Participants missing data in any of the variables were excluded from descriptive statistical analysis.
Abbreviations: MBI, mild behavioral impairment; CN, cognitively normal; MCI, mild cognitive impairment; SD, sleep disturbance

**Table 2.** Participant Characteristics for Analyses 1, 2, 3 and 4.

| **Analysis 1** | **Total (n=11277)** | **MBI (n=1562)** | **No MBI (n=9715)** | **p** |
| --- | --- | --- | --- | --- |
| **n** | 11277 | 1562 | 9715 |  |
| **First-Visit Age (Years)** | 72.7 (8.8), 50-104 | 72.8 (8.5), 50-104 | 72.7 (8.8), 50-101 | 0.501 <sup>w</sup> |
| **Females** | 6658 (59.0) | 705 (45.1) | 5953 (61.3) | <0.001 <sup>c</sup> |
| **Education (Years)** | 15.7 (3.1), 0-29 | 15.7 (3.2), 1-29 | 15.7 (3.0), 0-29 | 0.869 <sup>w</sup> |
| **Cognitive Diagnosis** |  |  |  | <0.001 <sup>c</sup> |
| CN | 8579 (76.1) | 768 (49.2) | 7811 (80.4) |  |
| MCI | 2698 (23.9) | 794 (50.8) | 1904 (19.6) |  |
| **Race: White\*** | 8721 (77.3) | 1301 (83.3) | 7420 (76.4) | <0.001 <sup>c</sup> |
| **Analysis 2** | **Total (n=10535)** | **SD Present (n=820)** | **SD Absent (n=9715)** | **p** |
| **First-Visit Age (Years)** | 72.7 (8.8), 50-101 | 72.2 (8.8), 50-96 | 72.7 (8.8), 50-101 | 0.106 <sup>w</sup> |
| **Females** | 6412 (60.9) | 459 (56.0) | 5953 (61.3) | 0.003 <sup>c</sup> |
| **Education (Years)** | 15.7 (3.2), 0-29 | 15.5 (3.2), 0-25 | 15.7 (3.0), 0-29 | 0.054 <sup>w</sup> |
| **Cognitive Diagnosis** |  |  |  | <0.001 <sup>c</sup> |
| CN | 8375 (79.5) | 564 (68.8) | 7811 (80.4) |  |
| MCI | 2160 (20.5) | 256 (31.2) | 1904 (19.6) |  |
| **Race: White\*** | 8063 (76.5) | 643 (78.4) | 7420 (76.4) | 0.115 <sup>c</sup> |
| **Analysis 3** | **Total (n=1567)** | **MBI Present (n=707)** | **MBI Absent (n=860)** | **p** |
| **First-Visit Age (Years)** | 72.2 (8.8), 50-101 | 72.2 (8.8), 50-101 | 72.3 (8.8), 50-96 | 0.877<sup>w</sup> |
| **Females** | 785 (50.1) | 307 (43.4) | 478 (55.6) | <0.001 <sup>c</sup> |
| **Education (Years)** | 15.4 (3.3), 0-30 | 15.1 (3.5), 0-30 | 15.6 (3.2), 0-25 | 0.017 <sup>w</sup> |
| **Cognitive Diagnosis** |  |  |  | <0.001 <sup>c</sup> |
| CN | 797 (50.9) | 233 (33.0) | 564 (65.6) |  |
| MCI | 770 (49.1) | 474 (67.0) | 296 (34.4) |  |
| **Race: White\*** | 1260 (80.4) | 581 (82.2) | 679 (79.0) | 0.008 <sup>c</sup> |
| **Analysis 4** | **Total (n=11921)** | **MBI Present (n=1902)** | **MBI Absent (n=10019)** | **p** |
| **First-Visit Age (Years)** | 72.9 (8.8), 50-104 | 73.1 (8.6), 50-104 | 72.8 (8.8), 50-101 | 0.089 <sup>w</sup> |
| **Females** | 6991 (58.6) | 871 (45.8) | 6120 (61.1) | <0.001 <sup>c</sup> |
| **Education (Years)** | 15.7 (3.1), 0-29 | 15.6 (3.2), 1-29 | 15.7 (3.0), 0-29 | 0.218 <sup>w</sup> |
| **Cognitive Diagnosis** |  |  |  | <0.001 <sup>c</sup> |
| CN | 8628 (72.4) | 783 (41.2) | 7845 (78.3) |  |
| MCI | 3293 (27.6) | 1119 (58.8) | 2174 (21.7) |  |
| **Race: White\*\*** | 9269 (77.8) | 1590 (83.6) | 7679 (76.6) | <0.001 <sup>c</sup> |

*Note:* All values for continuous variables are shown in mean (standard deviation). Values were rounded to one decimal place where appropriate, except for p-values which were rounded to three decimal places. Categorical variables are shown as n (%). Participants missing data for any of the variables were excluded from descriptive statistical analysis. Analysis (1): baseline mild behavioral impairment (MBI) and incident sleep disturbance (SD); (2) baseline SD and incident MBI; (3) baseline MBI in those with SD and incident dementia; (4) baseline MBI in those without SD and incident dementia
\*Non-white races: Black, First Nations American/Alaskan, Native Hawaiian/Other Pacific Islander, Asian or Other. w – Wilcoxon Rank Sum (Mann-Whitney) Test; c – Chi-Squared Test

### Analysis 1: Association Between Baseline MBI and Incident Sleep Disturbance

Of the total sample (*n*=11277), 6658 (59.0%) were female, with a mean first-visit age of 72.7 years (*SD=*8.8, range=50-104), and mean 15.7 (SD=3.1, range=0-29) years of education. Of this sample, 8579 (76.1%) were CN, 2698 (23.9%) had MCI, and 8721 (77.3%) were White. A total of 1562 (13.9%) participants had MBI, and 9715 (86.1%) had no MBI. The MBI group comprised significantly more males and those with MCI [Table 2].

Over the total follow-up period of 4.8 (SD=3.6, range=0.4-16.3) years, 2467 (21.9%) individuals developed SD before dementia. KM survival curves at a median follow-up time of 6 years showed that sleep-disturbance-free survival in the no MBI group was 75.5% (95%CI:74.3-76.6) and in the MBI group was 47.7% (95% CI:44.1-51.5) [Figure 2]. Cox proportional hazards regressions showed that the MBI group had a 3.0-fold (95%CI:2.8-3.3*, q*<0.001) greater rate of developing SD relative to those without MBI [Table 3].

**Figure 2.**
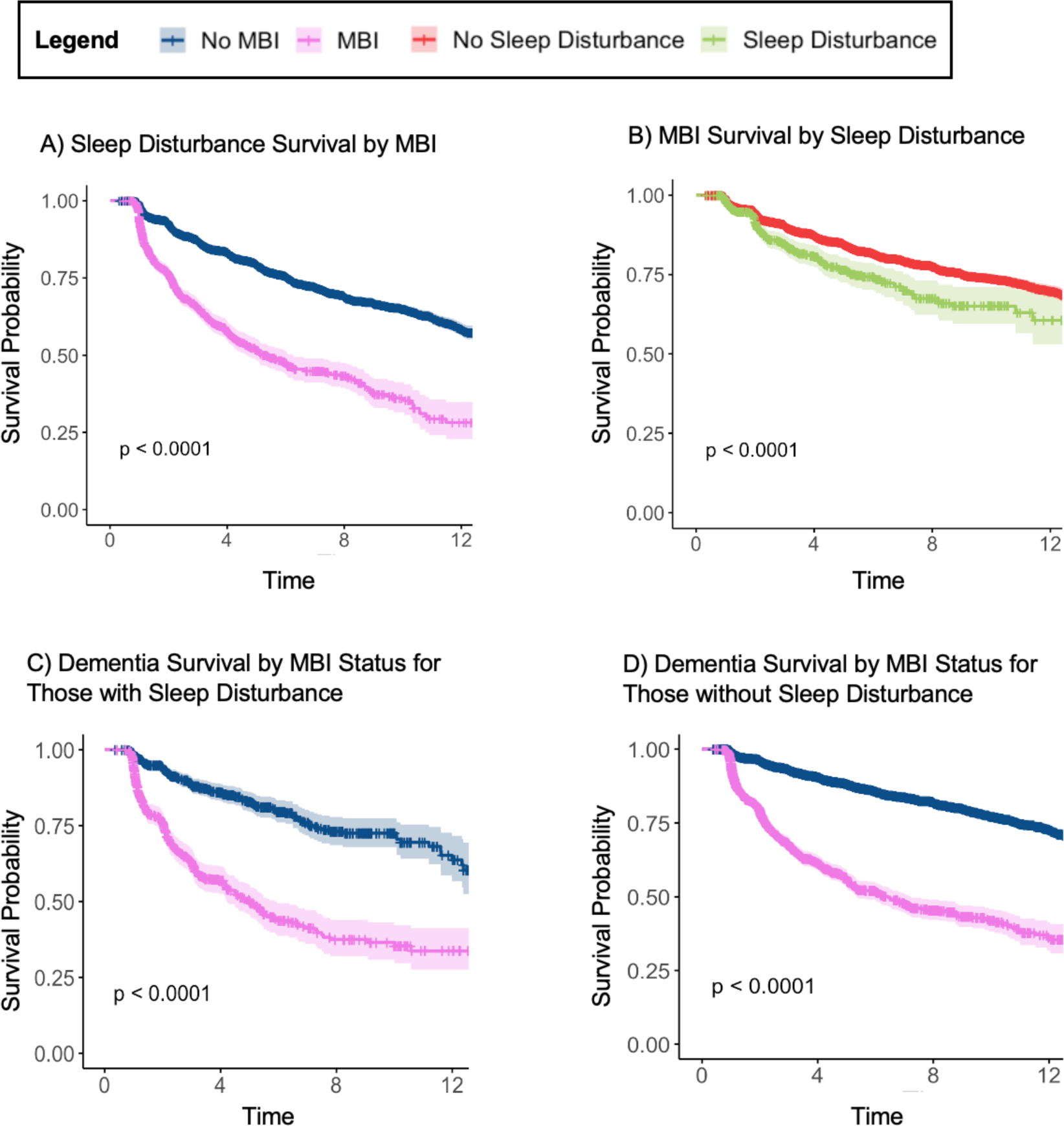
Kaplan-Meier Survival Curves. up to 12 years for (A) sleep disturbance-free survival based on MBI status (*n* = 11277); (B) MBI-free survival based on sleep disturbance (*n* = 10535); (C) dementia-free survival based MBI with concurrent sleep disturbance (*n* = 1567); (D) dementia-free survival based MBI without concurrent sleep disturbance (*n* =11921).

**Table 3.** Multivariable Cox regression models for the association between baseline MBI or sleep disturbance with incident sleep disturbance, MBI or dementia using MBI, SD and cognitive diagnosis as time-dependent variables.

| Exposure Variable | Outcome Variable | HR | 95% CI | *p* |
| --- | --- | --- | --- | --- |
| MBI | Sleep Disturbance | 3.04 | 2.77-3.33 | <0.001 |
| Sleep disturbance | MBI | 1.52 | 1.28-1.80 | <0.001 |
| MBI status in those with sleep disturbance | Dementia | 2.20 | 1.91-2.53 | <0.001 |
| MBI status in those without sleep disturbance | Dementia | 1.92 | 1.81-2.03 | <0.001 |

*Note.* Associations were tested using multivariable Cox regression models, adjusted for first-visit age, sex, years of education, cognitive diagnosis (CN/MCI), and race. Both MBI status and sleep disturbance were dichotomized. The false rate discovery method was used to adjust *p* values for multiple comparisons.
Abbreviations: MBI, mild behavioral impairment; CN, cognitively normal; MCI, mild cognitive impairment; HR, hazard ratio; CI, confidence interval

### Analysis 2: Association Between Baseline Sleep Disturbance and Incident MBI

Of the total sample (*n*=10535), 6412 (60.9%) were female, with a mean first-visit age of 72.7 (SD=8.8, range=50-101) years, and mean 15.7 (SD=3.2, range=0-29) years of education. Of this sample, 8375 (79.5%) were CN, 2160 (20.5%) had MCI, and 8063 (76.5%) were White. A total of 820 (7.8%) participants had SD, and 9715 (92.2%) had no SD. The SD group comprised significantly more females and CN individuals [Table 2].

Over the total follow-up period of 4.9 (SD=3.6, range=0.4-16.3), 1504 (14.3%) individuals developed MBI. KM survival curves at a median follow-up time of 6 years showed that MBI-free survival in the no SD group was 81.6% (95%CI:80.5-82.6) and in the SD group, was 74.3% (95%CI:70.2-78.7) [Figure 2]. Cox proportional hazards regressions showed that participants with SD at baseline had a 1.5-fold (95%CI:1.3-1.8, *q*<0.001) greater rate of developing MBI than those without SD [Table 3].

### Analysis 3: Association Between Baseline Sleep Disturbance and Incident Dementia, stratified by MBI status

Of the total sample (*n*=1567), 785 (50.1%) were female, with a mean first-visit age of 72.2 (SD=8.8, range=50-101) years and a mean 15.4 (SD=3.3, range=0-30) years of education. Of this sample, 797 (50.9%) were CN, 770 (49.1%) had MCI, and 1260 (80.4%) were White. Of the sample, 707 (45.1) individuals had MBI at baseline, and 860 (54.9) had no MBI at baseline. On average, participants with MBI were more likely to be male and have MCI [Table 2].

Over the average total follow-up period of 4.2 (SD=3.3, range=0.4-15.4) years, 411 (26.2%) individuals developed dementia. In those with SD, KM survival curves out to 6 years showed that dementia-free survival in the no MBI group was 79.6% (95% CI:76.0-83.4) and in the MBI group, was 43.7% (38.7-49.4) [Figure 2].

Cox proportional hazards regressions showed that sleep-disturbed older adults with MBI had a 2.2-fold (95%CI:1.9-2.6; q<0.001) greater rate of developing dementia compared to sleep-disturbed older adults without MBI [Table 3].

### Analysis 4: Association Between No Baseline Sleep Disturbance and Incident Dementia, stratified by MBI status

Of the total sample (*n*=11921), 6991 (50.1%) were female, with a mean first-visit age of 72.9 (SD=8.8, range=50-104) years and a mean 15.7 (SD=3.1, range=0-29) years of education. Of this sample, 8628 (72.4%) were CN, 3293 27.6%) had MCI, and 9269 (77.8%) were White. Of the sample, 1902 (16.0) individuals had MBI at baseline, and 10019 (84.0) had no MBI at baseline. On average, participants with MBI were more likely to be male and have MCI [Table 2].

Over the average total follow-up period of 5.0 (SD=3.6, range=0.4-16.3) years, 1959 (16.4%) individuals developed dementia. In those without SD, KM survival curves out to 6 years showed that dementia-free survival in the no MBI group was 85.6% (95% CI:84.7-86.5) and in the MBI group was 51.8% (48.8-54.9) [Figure 2].

Cox proportional hazards regressions showed that non-sleep-disturbed older adults with MBI had a 1.9-fold (95%CI:1.8-2.0; q<0.001) greater rate of developing dementia compared to non-sleep-disturbed older adults without MBI [Table 3].

## DISCUSSION

This study aimed to investigate the longitudinal associations between MBI and SD and explore the SD-associated risk for incident dementia in those with and without comorbid MBI. As hypothesized, this study demonstrated a bidirectional longitudinal relationship between MBI and SD; older adults with MBI at baseline developed SD at a greater rate than those without MBI, and those with SD at baseline developed MBI at a greater rate than those without SD. Furthermore, individuals concurrently reporting both SD and MBI and individuals experiencing only MBI developed dementia at faster rates than those with only SD or no SD, respectively.

This indicates that there may be additive effects of MBI and SD when considering the risk to dementia. Previous literature has cross-sectionally linked certain NPS to SD. One study found the presence of depression, disinhibition, and aberrant motor behavior to be associated with a greater frequency and severity of SD [43]. This finding may be explained by the difficulties these behavioral symptoms pose when attempting to fall asleep and stay asleep. Those experiencing depression have impaired sleep continuity with frequent and long periods of wakefulness, reduced sleep efficiency, delayed sleep onset, and a reduced total sleep time [9]. SD in behaviorally disturbed individuals may arise from a disruption of the homeostatic and circadian drives to sleep [44]. The reverse relationship has also been postulated, where SD may increase susceptibility to behavioral symptoms such as anxiety and depression [10], poor emotional generation and regulation [45], and decreased inhibition [11]. This association may be due to functional connectivity deficits between the amygdala and the ventral anterior cingulate cortex (vACC) [46], leading to enhanced amygdala responses to negative stimuli, causing increased aggression and agitation. Sleep deficiencies can also reduce the ability of the medial prefrontal cortex (mPFC) to inhibit amygdala activity, which, similarly, may lead to behavioral and emotional instability [47, 48]. Thus, in line with previous literature, the findings of the present study indicate a possible bidirectional relationship between SD and behavioral impairments, such that behavioral symptoms may lead to the development of SD and vice versa.

SD as a risk factor can contribute to underlying AD pathology [49–51] but may also be sequelae of existing AD pathology in older adults (Ju et al., 2014). Hence, the bidirectional association between MBI and SD and a higher rate of decline to dementia with both MBI and SD may be explained by a common pathology. Having MBI may indicate a more advanced pathology, which is more likely to disrupt regions of the brain that regulate sleep, leading to SD [16, 52–59]. Alternatively, a positive feedback mechanism may be in play such that increased SD contributes to MBI, and MBI contributes to SD. In such a scenario, therapies aimed at alleviating MBI symptoms may help reduce the development or exacerbation of SD, as the reduction of behavioral symptoms may increase the likelihood of falling asleep and staying asleep. Conversely, existing medications for the management of SD may help directly or indirectly reduce behavioral symptoms such as anxiety and depression [60].

Additionally, these results are consistent with previous studies that have established the association between MBI, sleep, and dementia [24, 26, 49]. First, MBI has been linked to incident cognitive decline and dementia [61]. The greater dementia risk in those with MBI may relate to the association of MBI with pathological hallmarks of AD, like the formation of amyloid-β (Aβ) plaques and hyperphosphorylation and aggregation of tau proteins [62]. Studies have shown MBI to be associated with Aβ accumulation [56, 63–65] and hyperphosphorylated tau deposition [66, 67]. Second, studies have found that in non-dementia older adults, those with SD have a greater risk of developing dementia (Lee et al., 2020; Robbins et al., 2021; Shi et al., 2018). Sleep-wake cycles influence Aβ levels in the brain, where decreased sleep leads to Aβ accumulation [49–51]. The formation of Aβ plaques is proposed to decrease sleep quality directly by disrupting neuronal function in brain regions involved in sleep-wake cycles [68] or indirectly, worsening the effects in a positive-feedback manner [49]. Studies have also found that tau accumulation increases with greater SD and vice versa [69]. In light of the association between MBI and SD with greater cognitive decline and incident dementia due to similar pathological changes, combining the markers may enhance the ability to detect or predict dementia onset.

Based on these findings, clinicians might consider screening for SD and further assess MBI presence to improve dementia risk assessment in their patients. Several studies suggest that combining markers of dementia, such as cognition, gait, and MBI, may improve dementia prognostication [70, 71]. Thus, these markers should be incorporated into dementia risk assessments to best aid early detection.

This study does have limitations. Using a single item from the NPI-Q to assess SD, rated by an informant, may not identify all presentations of SD, as it captures insomnia and excessive daytime sedation (EDS). While EDS is a symptom of a variety of SD [72], it is important to assess this relationship, specifically examining SD such as REM sleep disorder, sleep apnea, and restless leg syndrome. This will allow for a more complete understating of the impact of SD on incident dementia, especially as SD, such as REM sleep disorder, are associated with specific dementias like Lewy body dementia [73]. Additionally, the use of NPI-Q to derive MBI scores poses a potential limitation as this questionnaire was not designed initially to measure MBI. For instance, the two-consecutive visits method was used to operationalize MBI because the NPI-Q has a one-month reference range. However, because symptom presence between study visits outside of the reference range of the NPI-Q cannot be confirmed, the symptom persistence criteria required for MBI diagnosis might not be met. The MBI checklist (MBI-C) was specifically developed for the detection of MBI, has a 6-month reference range, and explicitly captures later-life emergent and persistent NPS [7, 74]. However, the MBI-C has only very recently been included in the NACC UDS as an optional measure, so it will take some time to generate sufficient data with this scale. Therefore, in legacy datasets, using operational definitions or algorithms to derive MBI status from other measures is necessary. Dichotomizing SD and MBI did not allow for the assessment of the severity of these items, which may have provided additional insights. Nonetheless, we did find significant associations with discrete variables. Future studies could explore this relationship with more robust and continuous indicators of MBI and SD to ascertain whether this relationship holds when the severity of symptoms is considered. Lastly, we acknowledge there are other variables, in addition to the covariates used in this study, that could impact the relationships investigated in this study. Therefore, future studies should investigate how other variables associated with increased dementia risk, such as obesity, type 2 diabetes, traumatic brain injury, stroke, and hearing loss, impact these relationships [75].

Regardless, this study establishes a starting point for further investigations regarding the bidirectional relationship between MBI and SD, the relationship between SD and dementia, and MBI and dementia. Additionally, by illustrating the bidirectional relationship and the relationship with incident dementia, this study emphasizes the importance of non-cognitive markers of dementia in pre-dementia screening, especially the importance of SD. Being longitudinal in design, this study allowed exploration of the bidirectional relationship between MBI and SD. The large sample size also provided sufficient statistical power to test several hypotheses, enabling a fuller understanding of the relationship between MBI, SD, and dementia.

## CONCLUSION

This study demonstrated that older adults with MBI have a greater rate of incident SD, and those with SD have a greater incidence of MBI. Furthermore, when participants experienced SD along with MBI, the rate of developing dementia was greater compared to experiencing only SD. These findings set the stage for treatment studies to determine if treating symptoms of MBI could reduce the incidence of SD and vice versa, as well as the impact of treating both on incident cognitive decline and dementia. Nonetheless, both sleep and MBI should be incorporated into dementia risk assessments to aid with early detection.

## ACKNOWLEDGEMENTS

The authors have no acknowledgments to report.

## FUNDING

- Dinithi Mudalige received funding from Alberta Innovates as part of the 2022 Summer Student Award and the Provincial Seniors Health and Continuing Care Studentship as part of the 2023 Summer Student Award; Alberta Innovates and Seniors Health played no role in any part of the study design, analysis, or interpretation of this study.
- Zahinoor Ismail is funded by the Canadian Institutes of Health Research (BCA2633) and the National Institute for Health and Care Research (NIHR) Exeter Biomedical Research Centre. The views expressed are those of the author(s) and not necessarily those of the NIHR or the Department of Health and Social Care.
- The NACC database is funded by NIA/NIH Grant U24 AG072122. NACC data are contributed by the NIA-funded ADRCs: P30 AG062429 (PI James Brewer, MD, PhD), P30 AG066468 (PI Oscar Lopez, MD), P30 AG062421 (PI Bradley Hyman, MD, PhD), P30 AG066509 (PI Thomas Grabowski, MD), P30 AG066514 (PI Mary Sano, PhD), P30 AG066530 (PI Helena Chui, MD), P30 AG066507 (PI Marilyn Albert, PhD), P30 AG066444 (PI John Morris, MD), P30 AG066518 (PI Jeffrey Kaye, MD), P30 AG066512 (PI Thomas Wisniewski, MD), P30 AG066462 (PI Scott Small, MD), P30 AG072979 (PI David Wolk, MD), P30 AG072972 (PI Charles DeCarli, MD), P30 AG072976 (PI Andrew Saykin, PsyD), P30 AG072975 (PI David Bennett, MD), P30 AG072978 (PI Neil Kowall, MD), P30 AG072977 (PI Robert Vassar, PhD), P30 AG066519 (PI Frank LaFerla, PhD), P30 AG062677 (PI Ronald Petersen, MD, PhD), P30 AG079280 (PI Eric Reiman, MD), P30 AG062422 (PI Gil Rabinovici, MD), P30 AG066511 (PI Allan Levey, MD, PhD), P30 AG072946 (PI Linda Van Eldik, PhD), P30 AG062715 (PI Sanjay Asthana, MD, FRCP), P30 AG072973 (PI Russell Swerdlow, MD), P30 AG066506 (PI Todd Golde, MD, PhD), P30 AG066508 (PI Stephen Strittmatter, MD, PhD), P30 AG066515 (PI Victor Henderson, MD, MS), P30 AG072947 (PI Suzanne Craft, PhD), P30 AG072931 (PI Henry Paulson, MD, PhD), P30 AG066546 (PI Sudha Seshadri, MD), P20 AG068024 (PI Erik Roberson, MD, PhD), P20 AG068053 (PI Justin Miller, PhD), P20 AG068077 (PI Gary Rosenberg, MD), P20 AG068082 (PI Angela Jefferson, PhD), P30 AG072958 (PI Heather Whitson, MD), P30 AG072959 (PI James Leverenz, MD).

## CONFLICT OF INTEREST

ZI reports serving as a consultant/advisor to Eisai, Lundbeck, Otsuka, and Roche. All authors have no conflict of interest to report.

## DATA AVAILABILITY

The data used in this study were obtained from NACC (https://naccdata.org/). Data are available free of charge to bona fide researchers who submit a research proposal.

## Notes

### Author Declarations

As determined by the University of Washington Human Subjects Division, use of the National Alzheimer's Coordinating Center (NACC) database itself is exempt from Institutional Review Board review. However, all contributing Alzheimer's Disease Research Centers are required to obtain informed consent from their participants and to maintain their own separate Institutional Review Board review and approval from their institution before submitting data to NACC.

## REFERENCES

[1] Lynch C (2020) World Alzheimer Report 2019: Attitudes to dementia, a global survey: Public health: Engaging people in ADRD research. Alzheimer’s & Dementia 16, e038255.

[2] World Health Organization (2021) Global status report on the public health response to dementia.

[3] Morris JC (1996) Classification of dementia and Alzheimer’s disease. Acta Neurologica Scandinavica 94, 41–50.

[4] Gauthier S, Albert M, Fox N, Goedert M, Kivipelto M, Mestre-Ferrandiz J, Middleton LT (2016) Why has therapy development for dementia failed in the last two decades? Alzheimer’s & Dementia 12, 60–64.

[5] Ismail Z, Black SE, Camicioli R, Chertkow H, Herrmann N, Laforce Jr R, Montero-Odasso M, Rockwood K, Rosa-Neto P, Seitz D (2020) Recommendations of the 5th Canadian Consensus Conference on the diagnosis and treatment of dementia. Alzheimer’s & Dementia 16, 1182–1195.

[6] Montero-Odasso M, Pieruccini-Faria F, Ismail Z, Li K, Lim A, Phillips N, Kamkar N, Sarquis-Adamson Y, Speechley M, Theou O, Verghese J, Wallace L, Camicioli R (2020) CCCDTD5 recommendations on early non cognitive markers of dementia: A Canadian consensus. Alzheimers Dement (N Y) 6, e12068.

[7] Ismail Z, Smith EE, Geda Y, Sultzer D, Brodaty H, Smith G, Agüera-Ortiz L, Sweet R, Miller D, Lyketsos CG (2016) Neuropsychiatric symptoms as early manifestations of emergent dementia: provisional diagnostic criteria for mild behavioral impairment. Alzheimer’s & Dementia 12, 195–202.

[8] Miner B, Kryger MH (2017) Sleep in the Aging Population. Sleep Med Clin 12, 31–38.

[9] Nutt D, Wilson S, Paterson L (2008) Sleep disorders as core symptoms of depression. Dialogues Clin Neurosci 10, 329–336.

[10] Babson KA, Trainor CD, Feldner MT, Blumenthal H (2010) A test of the effects of acute sleep deprivation on general and specific self-reported anxiety and depressive symptoms: An experimental extension. Journal of Behavior Therapy and Experimental Psychiatry 41, 297–303.

[11] Drummond SPA, Paulus MP, Tapert SF (2006) Effects of two nights sleep deprivation and two nights recovery sleep on response inhibition. Journal of Sleep Research 15, 261–265.

[12] Rouse HJ, Small BJ, Schinka JA, Loewenstein DA, Duara R, Potter H (2021) Mild behavioral impairment as a predictor of cognitive functioning in older adults. Int Psychogeriatr 33, 285–293.

[13] Kassam F, Chen H, Nosheny RL, McGirr A, Williams T, Ng N, Camacho M, Mackin RS, Weiner MW, Ismail Z (2022) Cognitive profile of people with mild behavioral impairment in Brain Health Registry participants. International Psychogeriatrics, 1–10.

[14] Creese B, Brooker H, Ismail Z, Wesnes KA, Hampshire A, Khan Z, Megalogeni M, Corbett A, Aarsland D, Ballard C (2019) Mild behavioral impairment as a marker of cognitive decline in cognitively normal older adults. The American Journal of Geriatric Psychiatry 27, 823–834.

[15] Ismail Z, McGirr A, Gill S, Hu S, Forkert ND, Smith EE (2021) Mild Behavioral Impairment and Subjective Cognitive Decline Predict Cognitive and Functional Decline. J Alzheimers Dis 80, 459–469.

[16] Ghahremani M, Nathan S, Smith EE, McGirr A, Goodyear B, Ismail Z (2023) Functional connectivity and mild behavioral impairment in dementia-free elderly. Alzheimer’s & Dementia: Translational Research & Clinical Interventions 9, e12371.

[17] Matsuoka T, Ismail Z, Narumoto J (2019) Prevalence of Mild Behavioral Impairment and Risk of Dementia in a Psychiatric Outpatient Clinic.

[18] Kan CN, Cano J, Zhao X, Ismail Z, Chen CL, Xu X (2022) Prevalence, Clinical Correlates, Cognitive Trajectories, and Dementia Risk Associated With Mild Behavioral Impairment in Asians. J Clin Psychiatry 83.

[19] Wolfova K, Creese B, Aarsland D, Ismail Z, Corbett A, Ballard C, Hampshire A, Cermakova P (2022) Gender/Sex Differences in the Association of Mild Behavioral Impairment with Cognitive Aging. J Alzheimers Dis 88, 345–355.

[20] Vellone D, Ghahremani M, Goodarzi Z, Forkert ND, Smith EE, Ismail Z (2022) Apathy and APOE in mild behavioral impairment, and risk for incident dementia. Alzheimers Dement (N Y) 8, e12370.

[21] Ebrahim IM, Ghahremani M, Camicioli R, Smith EE, Ismail Z (2023) Effects of race, baseline cognition, and APOE on the association of affective dysregulation with incident dementia: A longitudinal study of dementia-free older adults. Journal of Affective Disorders 332, 9–18.

[22] Ismail Z, Ghahremani M, Amlish Munir M, Fischer CE, Smith EE, Creese B (2023) A longitudinal study of late-life psychosis and incident dementia and the potential effects of race and cognition. Nature Mental Health 1, 273–283.

[23] McGirr A, Nathan S, Ghahremani M, Gill S, Smith EE, Ismail Z (2022) Progression to Dementia or Reversion to Normal Cognition in Mild Cognitive Impairment as a Function of Late-Onset Neuropsychiatric Symptoms. Neurology 98, e2132.

[24] Shi L, Chen S-J, Ma M-Y, Bao Y-P, Han Y, Wang Y-M, Shi J, Vitiello MV, Lu L (2018) Sleep disturbances increase the risk of dementia: a systematic review and meta-analysis. Sleep medicine reviews 40, 4–16.

[25] Robbins R, Quan SF, Weaver MD, Bormes G, Barger LK, Czeisler CA (2021) Examining sleep deficiency and disturbance and their risk for incident dementia and all-cause mortality in older adults across 5 years in the United States. Aging (Albany NY) 13, 3254–3268.

[26] Lee W, Gray SL, Barthold D, Maust DT, Marcum ZA (2020) Association Between Informant-Reported Sleep Disturbance and Incident Dementia: An Analysis of the National Alzheimer’s Coordinating Center Uniform Data Set. Journal of Applied Gerontology 41, 285–294.

[27] Sterniczuk R, Theou O, Rusak B, Rockwood K (2013) Sleep disturbance is associated with incident dementia and mortality. Current Alzheimer research 10, 767–775.

[28] Beekly DL, Ramos EM, van Belle G, Deitrich W, Clark AD, Jacka ME, Kukull WA (2004) The national Alzheimer’s coordinating center (NACC) database: an Alzheimer disease database. Alzheimer Disease & Associated Disorders 18, 270–277.

[29] Besser L, Kukull W, Knopman DS, Chui H, Galasko D, Weintraub S, Jicha G, Carlsson C, Burns J, Quinn J (2018) Version 3 of the National Alzheimer’s Coordinating Center’s uniform data set. Alzheimer disease and associated disorders.

[30] Morris JC, Weintraub S, Chui HC, Cummings J, DeCarli C, Ferris S, Foster NL, Galasko D, Graff-Radford N, Peskind ER (2006) The Uniform Data Set (UDS): clinical and cognitive variables and descriptive data from Alzheimer Disease Centers. Alzheimer Disease & Associated Disorders 20, 210–216.

[31] Weintraub S, Salmon D, Mercaldo N, Ferris S, Graff-Radford NR, Chui H, Cummings J, DeCarli C, Foster NL, Galasko D (2009) The Alzheimer’s disease centers’ uniform data set (UDS): The neuropsychological test battery. Alzheimer disease and associated disorders 23, 91.

[32] Bliwise DL, Mercaldo ND, Avidan AY, Boeve BF, Greer SA, Kukull WA (2011) Sleep disturbance in dementia with Lewy bodies and Alzheimer’s disease: a multicenter analysis. Dementia and geriatric cognitive disorders 31, 239–246.

[33] Bubu OM, Williams ET, Umasabor-Bubu OQ, Kaur SS, Turner AD, Blanc J, Cejudo JR, Mullins AE, Parekh A, Kam K (2021) Interactive Associations of Neuropsychiatry Inventory-Questionnaire Assessed Sleep Disturbance and Vascular Risk on Alzheimer’s Disease Stage Progression in Clinically Normal Older Adults. Frontiers in Aging Neuroscience 13.

[34] Burke SL, Hu T, Spadola CE, Burgess A, Li T, Cadet T (2018) Treatment of Sleep Disturbance May Reduce the Risk of Future Probable Alzheimer’s Disease. Journal of Aging and Health 31, 322–342.

[35] Burke SL, Hu T, Spadola CE, Li T, Naseh M, Burgess A, Cadet T (2018) Mild cognitive impairment: associations with sleep disturbance, apolipoprotein e4, and sleep medications. Sleep Medicine 52, 168–176.

[36] Kaufer DI, Cummings JL, Ketchel P, Smith V, MacMillan A, Shelley T, Lopez OL, DeKosky ST (2000) Validation of the NPI-Q, a brief clinical form of the Neuropsychiatric Inventory. The Journal of neuropsychiatry and clinical neurosciences 12, 233–239.

[37] Sheikh F, Ismail Z, Mortby ME, Barber P, Cieslak A, Fischer K, Granger R, Hogan DB, Mackie A, Maxwell CJ (2018) Prevalence of mild behavioral impairment in mild cognitive impairment and subjective cognitive decline, and its association with caregiver burden. International Psychogeriatrics 30, 233–244.

[38] Mortby ME, Ismail Z, Anstey KJ (2018) Prevalence estimates of mild behavioral impairment in a population-based sample of pre-dementia states and cognitively healthy older adults. Int Psychogeriatr 30, 221–232.

[39] Pocock SJ, Clayton TC, Altman DG (2002) Survival plots of time-to-event outcomes in clinical trials: good practice and pitfalls. Lancet 359, 1686–1689.

[40] Lee M, Whitsel E, Avery C, Hughes TM, Griswold ME, Sedaghat S, Gottesman RF, Mosley TH, Heiss G, Lutsey PL (2022) Variation in Population Attributable Fraction of Dementia Associated With Potentially Modifiable Risk Factors by Race and Ethnicity in the US. JAMA Network Open 5, e2219672–e2219672.

[41] Chen J-H, Lin K-P, Chen Y-C (2009) Risk factors for dementia. Journal of the Formosan Medical Association 108, 754–764.

[42] R Core Team (2022), ed. Computing RFfS, Austria.

[43] García-Alberca JM, Lara JP, Cruz B, Garrido V, Gris E, Barbancho MÁ (2013) Sleep disturbances in Alzheimer’s disease are associated with neuropsychiatric symptoms and antidementia treatment. The Journal of nervous and mental disease 201, 251–257.

[44] Borbély AA (1982) A two process model of sleep regulation. Hum neurobiol 1, 195–204.

[45] Palmer CA, Alfano CA (2017) Sleep and emotion regulation: An organizing, integrative review. Sleep Medicine Reviews 31, 6–16.

[46] Motomura Y, Kitamura S, Oba K, Terasawa Y, Enomoto M, Katayose Y, Hida A, Moriguchi Y, Higuchi S, Mishima K (2013) Sleep debt elicits negative emotional reaction through diminished amygdala-anterior cingulate functional connectivity. PloS one 8, e56578.

[47] Motomura Y, Katsunuma R, Yoshimura M, Mishima K (2017) Two days’ sleep debt causes mood decline during resting state via diminished amygdala-prefrontal connectivity. Sleep 40.

[48] Ismail Z, Herrmann N, Francis PL, Rothenburg LS, Lobaugh NJ, Leibovitch FS, Black SE, Lanctôt KL (2009) A SPECT Study of Sleep Disturbance and Alzheimer’s Disease. Dementia and Geriatric Cognitive Disorders 27, 254–259.

[49] Ju Y-ES, Lucey BP, Holtzman DM (2014) Sleep and Alzheimer disease pathology--a bidirectional relationship. Nature reviews. Neurology 10, 115–119.

[50] Winer JR, Mander BA, Kumar S, Reed M, Baker SL, Jagust WJ, Walker MP (2020) Sleep Disturbance Forecasts β-Amyloid Accumulation across Subsequent Years. Current Biology 30, 4291–4298.e4293.

[51] Kang J-E, Lim MM, Bateman RJ, Lee JJ, Smyth LP, Cirrito JR, Fujiki N, Nishino S, Holtzman DM (2009) Amyloid-β dynamics are regulated by orexin and the sleep-wake cycle. Science 326, 1005–1007.

[52] Matsuoka T, Imai A, Narumoto J (2023) Neuroimaging of mild behavioral impairment: A systematic review. Psychiatry and Clinical Neurosciences Reports 2, e81.

[53] Matuskova V, Ismail Z, Nikolai T, Markova H, Cechova K, Nedelska Z, Laczó J, Wang M, Hort J, Vyhnalek M (2021) Mild Behavioral Impairment Is Associated With Atrophy of Entorhinal Cortex and Hippocampus in a Memory Clinic Cohort. Front Aging Neurosci 13, 643271.

[54] Creese B, Arathimos R, Brooker H, Aarsland D, Corbett A, Lewis C, Ballard C, Ismail Z (2021) Genetic risk for Alzheimer’s disease, cognition, and mild behavioral impairment in healthy older adults. *Alzheimer’s & Dementia: Diagnosis*, Assessment & Disease Monitoring 13, e12164.

[55] Miao R, Chen HY, Robert P, Smith EE, Ismail Z (2021) White matter hyperintensities and mild behavioral impairment: Findings from the MEMENTO cohort study. Cereb Circ Cogn Behav 2, 100028.

[56] Miao R, Chen HY, Gill S, Naude J, Smith EE, Ismail Z (2022) Plasma β-Amyloid in Mild Behavioural Impairment - Neuropsychiatric Symptoms on the Alzheimer’s Continuum. J Geriatr Psychiatry Neurol 35, 434–441.

[57] Naude JP, Gill S, Hu S, McGirr A, Forkert ND, Monchi O, Stys PK, Smith EE, Ismail Z, Initiative AsDN (2020) Plasma neurofilament light: a marker of neurodegeneration in mild behavioral impairment. Journal of Alzheimer’s disease 76, 1017–1027.

[58] Shu J, Qiang Q, Yan Y, Wen Y, Ren Y, Wei W, Zhang L (2021) Distinct patterns of brain atrophy associated with mild behavioral impairment in cognitively normal elderly adults. International Journal of Medical Sciences 18, 2950.

[59] Gill S, Wang M, Mouches P, Rajashekar D, Sajobi T, MacMaster FP, Smith EE, Forkert ND, Ismail Z (2021) Neural correlates of the impulse dyscontrol domain of mild behavioral impairment. Int J Geriatr Psychiatry 36, 1398–1406.

[60] Krystal AD (2020) Sleep therapeutics and neuropsychiatric illness. Neuropsychopharmacology 45, 166–175.

[61] Creese B, Ismail Z (2022) Mild behavioral impairment: measurement and clinical correlates of a novel marker of preclinical Alzheimer’s disease. Alzheimers Res Ther 14, 2.

[62] Jack Jr CR, Knopman DS, Jagust WJ, Petersen RC, Weiner MW, Aisen PS, Shaw LM, Vemuri P, Wiste HJ, Weigand SD (2013) Update on hypothetical model of Alzheimer’s disease biomarkers. Lancet neurology 12, 207.

[63] Sun Y, Xu W, Chen K-L, Shen X-N, Tan L, Yu J-T, for the Alzheimer’s Disease Neuroimaging I (2021) Mild behavioral impairment correlates of cognitive impairments in older adults without dementia: mediation by amyloid pathology. Translational Psychiatry 11, 577.

[64] Lussier FZ, Pascoal TA, Chamoun M, Therriault J, Tissot C, Savard M, Kang MS, Mathotaarachchi S, Benedet AL, Parsons M, Qureshi MNI, Thomas É M, Shin M, Dion LA, Massarweh G, Soucy JP, Tsai IH, Vitali P, Ismail Z, Rosa-Neto P, Gauthier S (2020) Mild behavioral impairment is associated with β-amyloid but not tau or neurodegeneration in cognitively intact elderly individuals. Alzheimers Dement 16, 192–199.

[65] Ismail Z, Leon R, Creese B, Ballard C, Robert P, Smith EE (2023) Optimizing detection of Alzheimer’s disease in mild cognitive impairment: a 4-year biomarker study of mild behavioral impairment in ADNI and MEMENTO. Mol Neurodegener **in press**.

[66] Johansson M, Stomrud E, Insel PS, Leuzy A, Johansson PM, Smith R, Ismail Z, Janelidze S, Palmqvist S, van Westen D, Mattsson-Carlgren N, Hansson O (2021) Mild behavioral impairment and its relation to tau pathology in preclinical Alzheimer’s disease. Transl Psychiatry 11, 76.

[67] Ghahremani M, Wang M, Chen H-Y, Zetterberg H, Smith E, Ismail Z, Initiative AsDN (2023) Plasma Phosphorylated Tau at Threonine 181 and Neuropsychiatric Symptoms in Preclinical and Prodromal Alzheimer Disease. Neurology 100, e683–e693.

[68] Braak H, Braak E (1991) Neuropathological stageing of Alzheimer-related changes. Acta neuropathologica 82, 239–259.

[69] Wang C, Holtzman DM (2020) Bidirectional relationship between sleep and Alzheimer’s disease: role of amyloid, tau, and other factors. Neuropsychopharmacology 45, 104–120.

[70] Verghese J, Wang C, Lipton RB, Holtzer R (2013) Motoric cognitive risk syndrome and the risk of dementia. J Gerontol A Biol Sci Med Sci 68, 412–418.

[71] Guan DX, Chen H-Y, Camicioli R, Montero-Odasso M, Smith EE, Ismail Z (2022) Dual-task gait and mild behavioral impairment: The interface between non-cognitive dementia markers. Experimental Gerontology 162, 111743.

[72] Brown J, Makker HK (2020) An approach to excessive daytime sleepiness in adults. BMJ 368.

[73] Boeve BF (2019) REM sleep behavior disorder associated with dementia with Lewy bodies. Rapid-eye-movement sleep behavior disorder, 67-76.

[74] Hu S, Patten S, Charlton A, Fischer K, Fick G, Smith EE, Ismail Z (2023) Validating the Mild Behavioral Impairment Checklist in a Cognitive Clinic: Comparisons With the Neuropsychiatric Inventory Questionnaire. J Geriatr Psychiatry Neurol 36, 107–120.

[75] Livingston G, Huntley J, Sommerlad A, Ames D, Ballard C, Banerjee S, Brayne C, Burns A, Cohen-Mansfield J, Cooper C (2020) Dementia prevention, intervention, and care: 2020 report of the Lancet Commission. The Lancet 396, 413–446.

